# Cardiac Output Monitoring via an Automatic Arm Cuff Device: Potential in Surgical Patients

**DOI:** 10.1101/2025.10.09.25337689

**Authors:** Mahdi Jazini, Hadi Daher, Ravinder Kumar, Vishaal Dhamotharan, Abhijeet M. Sathe, Yaroslava Longhitano, Ezeldeen Abuelkasem, Anand Chandrasekhar, Michael R. Pinsky, Kathirvel Subramaniam, Raymond M. Planinsic, Jin-Oh Hahn, Sanjeev G. Shroff, Kimberly Howard-Quijano, Aman Mahajan, Ramakrishna Mukkamala

**Affiliations:** Department of Bioengineering, University of Pittsburgh, Pittsburgh, PA, USA; Department of Anesthesiology and Perioperative Medicine, University of Pittsburgh School of Medicine, Pittsburgh, PA, USA; Department of Critical Care Medicine, University of Pittsburgh School of Medicine, Pittsburgh, PA, USA; Department of Mechanical Engineering, University of Maryland, College Park, MD, USA

**Author notes:** Equally contributing first authors (Mahdi Jazini, Hadi Daher, Ravinder Kumar). Submitting author contact information: Ramakrishna Mukkamala, PhD, Leighton E. and Mary N. Orr Professor, University of Pittsburgh, 408 Benedum Hall, 3700 O’Hara Street, Pittsburgh, PA 15261.

## Abstract

Pulse contour devices estimate cardiac output (CO) by analysis of an invasive blood pressure (BP) waveform. However, arterial catheters are only employed in the highest risk patients. By contrast, automatic arm cuff devices are used for non-invasive BP monitoring in numerous hospital patients. Furthermore, these devices directly measure cuff pressure oscillations, which are directly related to pulsatile blood volume rather than BP. The hypothesis of this study was that automatic arm cuff devices can offer an effective means to monitor CO. A custom-built cuff device was employed to measure cuff pressure and make a unique measurement of the volume of air pumped into and out of the cuff during linear deflation. Cuff device measurements, an invasive BP waveform via a radial artery catheter, and reference CO via pulmonary artery catheter-bolus thermodilution were obtained during major interventions from 24 liver transplant and 10 cardiac surgery patients. A basic analysis was applied to estimate CO in L/min as the product of heart rate, the maximum cuff pressure oscillation amplitude, the compliance of the cuff-arm system derived from the measured cuff volume-pressure relationship, and the body to arm surface area ratio derived using anthropometric information. This arm cuff-based method yielded a correlation of 0.60 and concordance rate of 83% against the reference CO. For comparison, classic pulse contour analysis of the invasive BP waveform produced a correlation of 0.62 and concordance rate of 81%. These initial findings indicate the potential for CO monitoring via a non-invasive automatic arm cuff device in surgical patients.

## Introduction

Pulse contour devices estimate cardiac output (CO) by analysis of a blood pressure (BP) waveform. These devices offer seamless, continuous CO monitoring in patients with indication for radial artery catheterization and are used in an appreciable fraction of these patients today^1,2^. However, the devices must approximate the arterial compliance scale factor, which relates BP waveform features (e.g., pulse pressure (PP) times heart rate (HR), diastolic decay time constant) to CO in L/min, from the BP levels and patient anthropometric information (e.g., sex, age, height, weight) using a formula built via a training dataset with reference CO measurements^3^. These devices may therefore be better suited for tracking CO changes in a patient (“CO trending”). Furthermore, arterial catheters are invasive and only employed in the highest risk patients. Pulse contour analysis may also be applied to the finger BP waveform obtained with volume-clamp devices for non-invasive and continuous CO monitoring. However, volume-clamp devices employ a special, servo-controlled finger cuff embedded with an optical sensor and may be vulnerable to common episodes of finger hypoperfusion^4^. As a result, these devices are rarely used in our hospital and have limited uptake elsewhere^1^.

By contrast, automatic arm cuff devices are employed for non-invasive BP monitoring in numerous hospital patients and even in those with arterial catheters in place. These devices estimate systolic and diastolic BP (SP and DP) from the measured cuff pressure oscillations and applied cuff pressure. The raw cuff pressure oscillation waveform is related to pulsatile blood volume oscillations and may therefore reflect blood flow rate more directly than a BP waveform reflects blood flow rate. Furthermore, the brachial artery measurement site is proximal to the radial and finger arteries and is therefore a better indicator of heart performance while being robust to hypoperfusion. Although automatic arm cuff devices do not permit continuous monitoring, they can and are used for frequent measurements in surgical and intensive care patients (e.g., every 3-5 min)^1^. Note that measurements at higher frequency may not be clinically actionable anyhow. For example, pulse contour devices are continuous yet often used only to measure 5-min averages of CO^5^. Automatic arm cuff devices can also be readily used for on-demand measurement (e.g., during interventions) by a mere press of a button.

Our hypothesis is that automatic arm cuff devices can offer an effective means to monitor CO. To initially test this hypothesis, we recorded measurements with a custom-built cuff device, the invasive radial BP waveform, and reference CO via pulmonary artery catheter-bolus thermodilution in surgical patients and estimated CO from the cuff device measurements using a basic analysis without the need for any training data. We found, perhaps surprisingly, that this arm cuff-based method was similar in CO measurement accuracy to classic invasive pulse contour analysis.

## Methods

### Custom Cuff Device

We used a custom-built automatic arm cuff device in this study. We first provide rationale for this custom-built “oscillometric” device and then briefly present the physical device along with exemplary measurements below.

#### Rationale

Conventionally, oscillometric devices rapidly inflate the cuff wrapped around the upper arm to a supra-systolic level and then slowly deflate the cuff to a sub-diastolic level while measuring the air pressure inside the cuff. The applied cuff pressure is superimposed with small oscillations indicating the pulsatile blood volume in the brachial artery (e.g., during systole, blood volume↑ → cuff volume↓ → cuff pressure↑ due to Boyle’s law). Because the relationship between transmural pressure (i.e., internal BP – external cuff pressure) and blood volume in arteries is sigmoidal with linear range around zero transmural pressure, the amplitude of the cuff pressure oscillations rises to a maximum when the applied cuff pressure is near the mean BP and then falls with the slowly decreasing cuff pressure. BP is estimated from the cuff pressure oscillation amplitude versus applied cuff pressure function (inverted U shaped “oscillogram”)^6^.

However, the cuff pressure-volume relationship, which approximately transduces the blood volume oscillations to cuff pressure oscillations^7^, changes with the cuff size, the way the cuff is wrapped around the arm, and the characteristics of the arm. This variable transducer effect can cause the cuff pressure oscillation amplitude to vary even without a change in the blood volume oscillations. For illustrative purposes, we interfaced a home device to an external pressure sensor and a non-invasive BP (NIBP) simulator with the maximum volume oscillation fixed and wrapped the cuff around a rigid mandrel with and without foam covering. Fig. 1A shows the resulting cuff pressure oscillations versus the applied cuff pressure. The cuff pressure oscillations for the cuff-rigid mandrel system were larger than for the cuff-rigid-foam mandrel system despite the same maximum volume oscillation. We then injected a known amount of air volume into each cuff-mandrel system in steps with a syringe and measured the steady-state cuff pressure with the external pressure sensor. Fig. 1B shows the resulting cuff pressure-volume relationships. The cuff-rigid mandrel system yielded larger cuff pressure oscillations due to the higher elastance (lower compliance) of the system. We therefore incorporated a unique measurement of the volume of air pumped into and out of the cuff along with the traditional cuff pressure measurement to determine the compliance of the cuff-arm system and thereby convert the cuff pressure oscillations in mmHg to blood volume oscillations in ml.

**Fig. 1:**
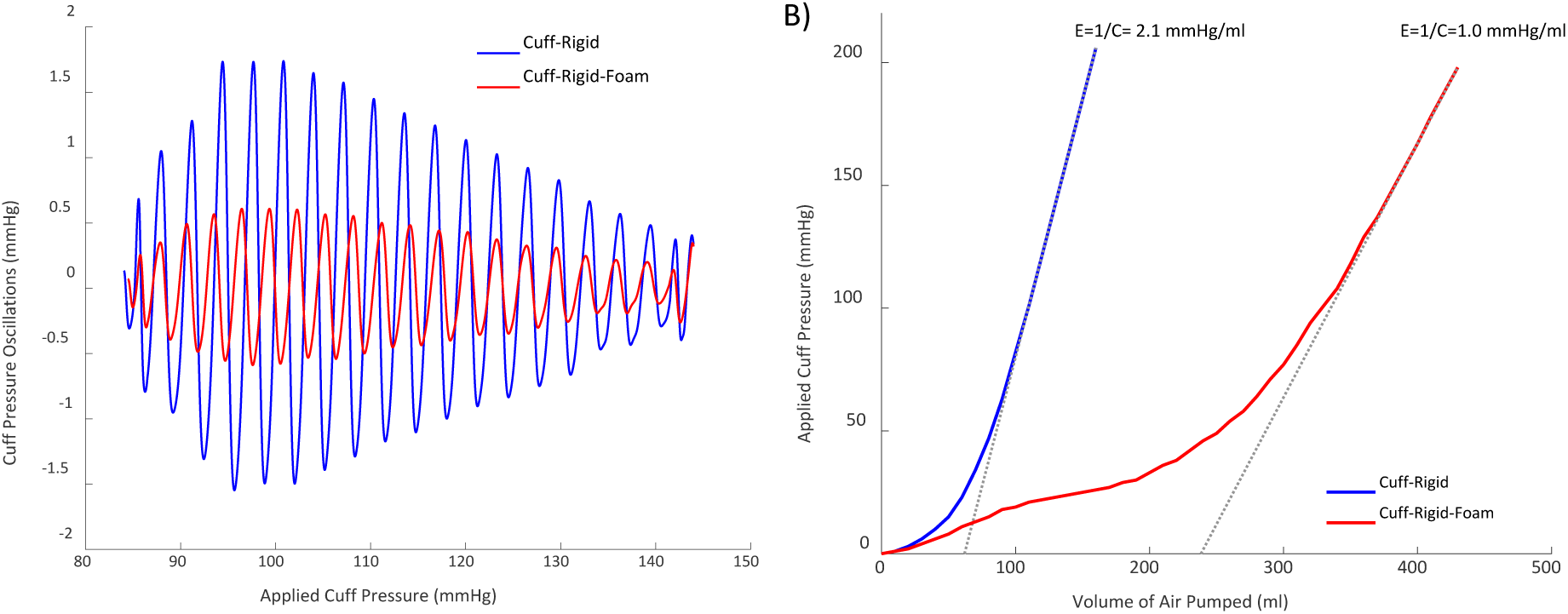
Rationale for measuring the volume of air pumped into and out of the cuff in addition to traditional cuff pressure with an automatic arm cuff device. **(A)** Cuff pressure oscillations versus applied cuff pressure obtained with a home device (BP7100 3 Series, Omron Healthcare) interfaced to an external pressure sensor (DPM2Plus Universal Pressure Meter Tester, Fluke Biomedical) and an NIBP simulator (ProSim 8, Fluke Biomedical) at fixed simulator settings (120/80 mmHg, 90 bpm, and 0.65 ml) and with the cuff wrapped around a rigid mandrel with and without polyethylene foam covering. NIBP is non-invasive blood pressure. The cuff-rigid mandrel system yielded larger cuff pressure oscillations than the cuff- rigid-foam mandrel system despite the same maximum volume oscillation setting. **(B)** Steady- state cuff pressure-volume relationships obtained with a syringe to inject known air volume into each cuff-mandrel system and with the external pressure sensor. The cuff-rigid mandrel system yielded large cuff pressure oscillations because of its higher elastance (E)/lower compliance (C). An additional measurement of the volume of air pumped into and out of the cuff can be used to determine the compliance of the cuff-arm system to convert the measured cuff pressure oscillations in mmHg to blood volume oscillations in ml and thereby facilitate cardiac output (CO) estimation.

Since the blood volume oscillations vary with the applied cuff pressure, conventional pulse contour analysis may not be directly translatable to the measured cuff pressure oscillation waveform. We therefore incorporated a period of constant cuff pressure, similar to central BP cuff devices^8^, to obtain a clean and steady waveform (“pulse volume recording (PVR)”) that could facilitate CO estimation.

We set the cuff inflation/deflation pattern by considering how existing cuff devices operate and available patient data. The devices that we studied (Carescape B850, GE; Intellivue, Philips; BP7100 3 Series, Omron; and WatchBP Office, Microlife) operate via cuff deflation at a rate of 3-4.5 mmHg/sec. A deflation rate of 4 mmHg/sec may be suitable for HR > 40 bpm (BP7100 3 Series). In public surgical and intensive care patient databases (PulseDB)^9^, 95% of the systolic BP values were < 150 mmHg; the average of the mean BP values was 82 mmHg; and 5% of the HR values were < 53 bpm. We employed linear deflation at a rate of 4 mmHg/sec from a maximum cuff pressure of 170 mmHg (or 200 mmHg for patients with high BP) to 10-20 mmHg to obtain well sampled and complete oscillograms for all measurements. We further set the subsequent cuff pressure to a constant level of 80 mmHg for 30 sec to yield a large amplitude PVR on average. We repeated this inflation/deflation cycle after 1 min for averaging purposes.

Note that the existing devices, by contrast, detect the cuff pressure oscillation amplitude in real time to minimize the measurement time. The hospital devices (Carescape B850 and Intellivue) also employ step deflation, which does not yield as well sampled oscillograms as linear deflation but is more robust to artifact. These devices appear to leverage a recent measurement when available to employ fewer steps for further reducing the measurement time.

#### Physical Device and Exemplary Measurements

Fig. 2 illustrates the custom cuff device along with a block diagram. The main components are (i) an air pump to inflate the cuff; (ii) a valve to deflate the cuff; (iii) a pressure sensor to measure the cuff pressure; (iv) a bidirectional airflow (micro-electromechanical systems thermal mass flow) sensor to measure the volume of air pumped into and out of the cuff; and (v) a microprocessor to control the pump and valve and obtain the measurements. The pressure sensor was calibrated in both static and dynamic modes using a reference manometer (DPM2Plus Universal Pressure Meter Tester, Fluke Biomedical), and the airflow sensor was calibrated in both directions with a syringe of known air volume. An electromagnetic interference (EMI) shield covers the pressure sensor and its associated circuitry. The pump is open-loop controlled for rapid inflation, whereas the valve is closed-loop controlled for linear deflation at a 4 mmHg/sec rate. Key features of the device include quick-disconnect fittings for attaching hospital-grade cuffs of all sizes; connectors for analog output of the two measurements; a manual knob for adjusting the maximum cuff pressure; an LCD for displaying the applied cuff pressure in real-time; LEDs for indicating device status; a switch for toggling between power from an internal rechargeable battery (> 30 hours of continuous operation) and an external adaptor; and a power button for safe operation. We verified the device with all cuff sizes using the NIBP simulator (ProSim 8, Fluke Biomedical) and for safety in the OR and ICU.

**Fig. 2:**
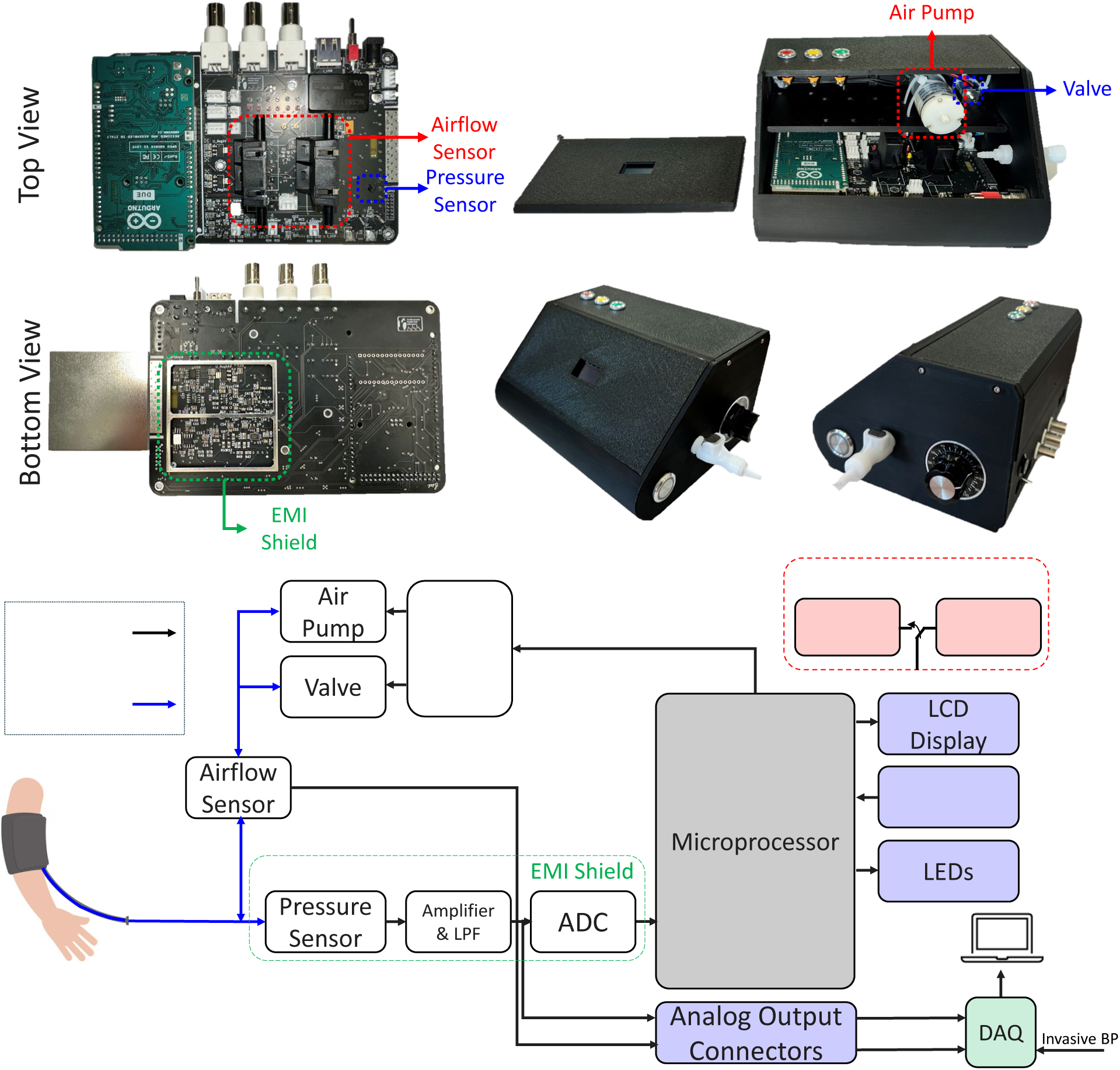
Custom automatic arm cuff device. Physical device along with a block diagram of the components connected via wires or tubes. EMI is electromagnetic interference; LPF, lowpass filter; ADC, analog-to-digital converter; DAQ, data acquisition unit.

Fig. 3 shows exemplary raw measurements of cuff pressure, cuff pressure oscillations, and the volume of air pumped into and out of the cuff (integral of airflow) with the device and two different cuff sizes from pilot patient studies. The device generally produced high-fidelity cuff pressure oscillations in absence of motion (Fig. 3BC), and the larger cuffs required more time (Fig. 3A) and airflow (Fig. 3D) for inflation to reach a fixed maximum applied cuff pressure.

**Fig. 3:**
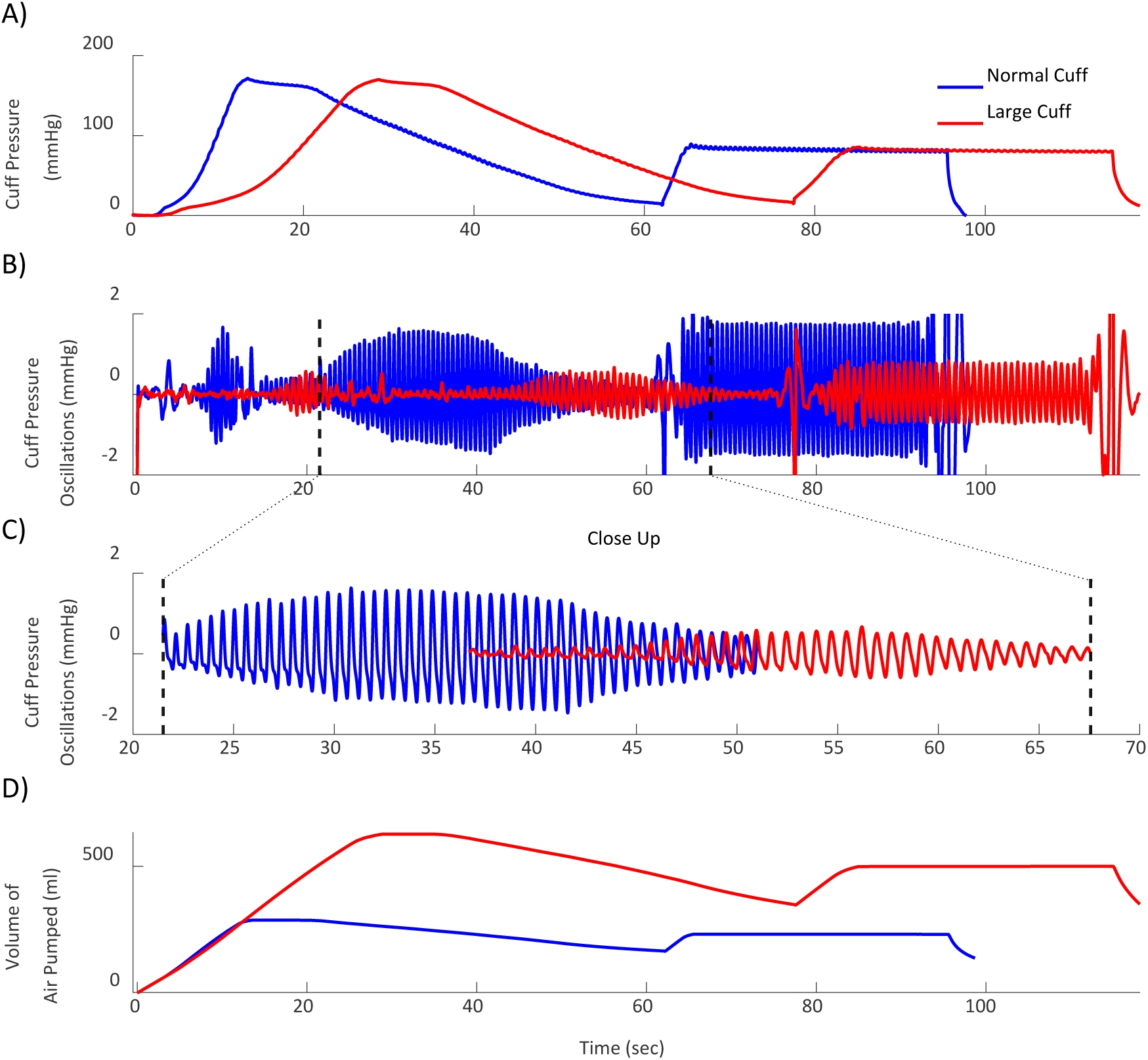
Exemplary custom cuff device measurements. Raw cuff pressure, cuff pressure oscillations via bandpass filtered cuff pressure, and volume of air pumped into and out of the cuff via the integral of airflow from pilot patients with different cuff sizes.

### Data Collection

We studied patients undergoing liver transplantation or cardiac surgery (on-pump coronary artery bypass grafting) at our hospital under IRB approval (University of Pittsburgh STUDY21110091) and with written, informed consent from the patients. The inclusion criteria were adults with pulmonary and radial artery catheters in place as part of routine care. The exclusion criteria were mechanical cardiac support (which distorts arterial waveforms) or significant tricuspid regurgitation (which is a contraindication for the reference thermodilution CO measurement) or significant aortic regurgitation (which is a contraindication of pulse contour analysis). We recruited a total of 37 patients for the study. This number of patients is not backed by a sample size calculation, because we had no preliminary data in support of the novel study hypothesis, but is comparable to previous, related studies^10–13^.

We began the study in the preoperative room by measuring the arm circumference of the patient. We wrapped an appropriately sized hospital-grade cuff (CRITIKON SOFT-CUF, GE) using the standard two finger approach on the arm opposite to the arm with the radial artery catheter to be used for pulse contour analysis. We also measured inter-arm BP differences in the liver transplant patients using the clinical NIBP cuff device. None of the patients showed inter-arm BP differences >10 mmHg.

We confirmed proper cuff wrapping and connected the cuff to the custom device in the OR. We also positioned a medical toboggan to protect the arm with the cuff of the cardiac surgery patients during the clinical procedures. We then interfaced the cuff device to a laptop via a data acquisition unit (USB-6002, National Instruments). We connected the bedside monitor (B850, GE; Canvas 1000, GE; or Intellivue, Philips), which acquired the clinical measurements including the invasive BP waveform via the radial artery catheter and the bolus thermodilution CO via the pulmonary artery catheter, to the same laptop via the USB and RS232 port. To synchronize all measurements, we connected the invasive BP analog output of the bedside monitor to the data acquisition unit for the cuff device. We used commercial software (DAQExpress, National Instruments) to control the acquisition of the cuff device and invasive BP waveform measurements and the freely available VitalRecorder software^14^ to control the acquisition of the bedside monitor data on the laptop. Fig. 4A illustrates the research and clinical devices and measurements and data acquisition instruments.

**Fig. 4:**
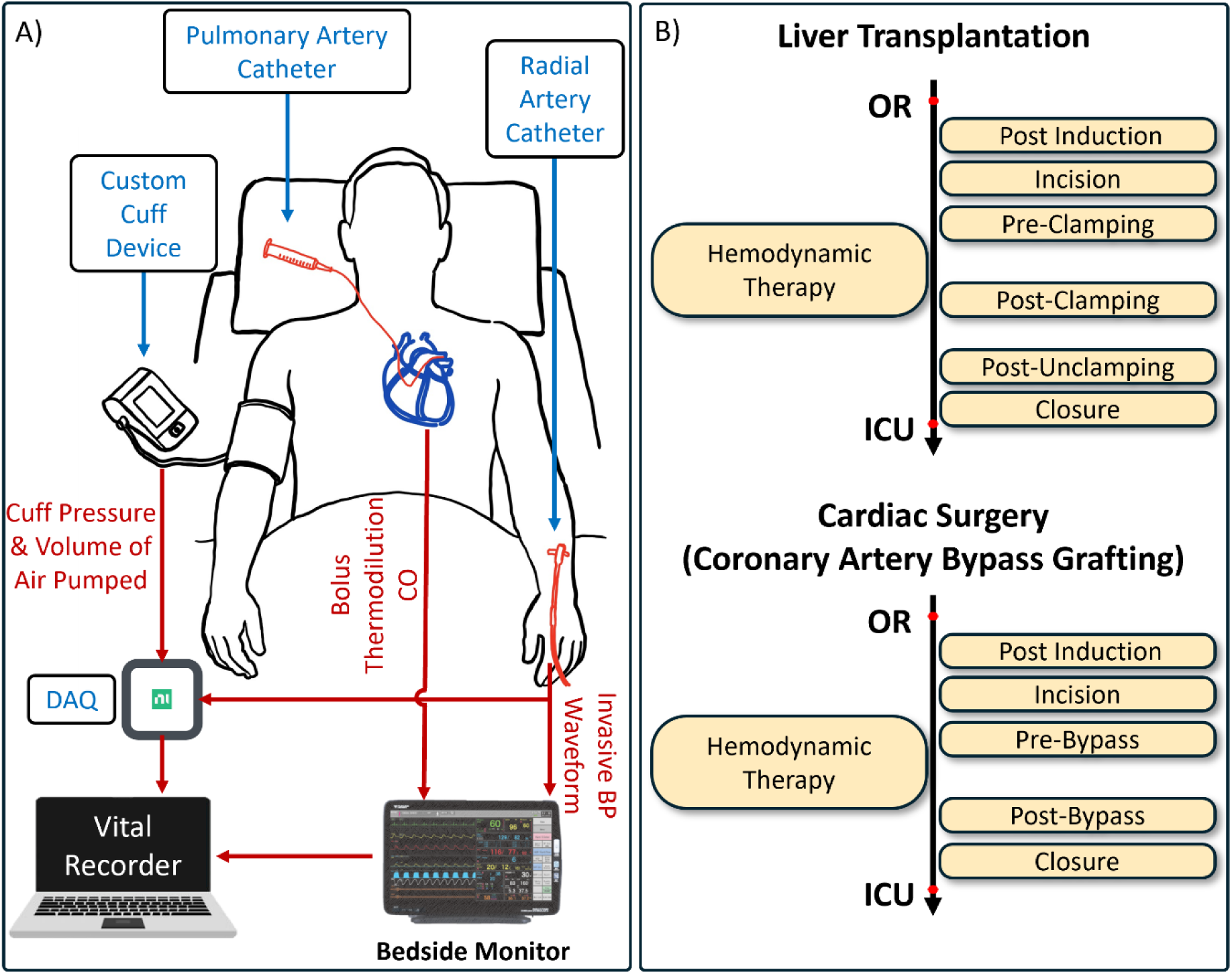
Patient data for analysis. **(A)** Main research and clinical devices and measurements and data acquisition instruments. **(B)** Measurement sets (cuff device measurements, invasive BP waveform, and reference CO) selected at hemodynamic therapy and indicated major surgical landmarks (when available) for analysis.

We simultaneously obtained cuff device measurements and bolus thermodilution CO in triplicate or quadruplicate using 10 ml 0.9% normal saline at room temperature at various time points including interventions during the surgery and the acute postoperative period in the ICU. We ensured that the radial artery catheter was regularly flushed (e.g., hourly) with the transducer kept at heart level. Throughout the study, we also entered time-stamped annotations in the VitalRecorder software.

We selected the measurement sets (cuff device measurements, invasive BP waveform, and reference CO) at major surgical landmarks, following any hemodynamic therapy, and after transfer to the ICU, as shown in Fig. 4B, to yield substantive reference CO changes in consecutive measurements for meaningful investigation of CO trending. We visually inspected the cuff device measurements and invasive BP waveforms, while blinded to the reference CO, and excluded measurement sets when either was contaminated by significant artifact for benchmarking purposes. This process reduced the study cohort to 34 patients.

### Data Analysis

We applied basic analyses to the cuff device measurements to estimate CO in two ways. Fig. 5 illustrates the analyses, which consists of three major steps. In the first step, the cuff pressure measurement was bandpass filtered (0.5 to 5 Hz) to obtain the cuff pressure oscillations and lowpass filtered (0.5 Hz) to obtain the applied cuff pressure. The peak-to-peak amplitude of the cuff pressure oscillations was then detected. The amplitudes were plotted against the applied cuff pressure during the deflation, and the resulting oscillogram was smoothed using a five-beat moving average. The maximum amplitude and corresponding HR were extracted from the oscillogram and multiplied together (ΔO_max_·HR) for a first surrogate of the arm blood flow rate in mmHg/min. The average of the amplitude and HR during the PVR were extracted and multiplied together (ΔPVR·HR) for a second surrogate of the arm blood flow rate in mmHg/min. In the second step, the volume of air pumped into and out of the cuff (integral of airflow) was plotted against the applied cuff pressure. Although this cuff volume-pressure relationship showed a hysteresis loop over the inflation and deflation periods, the compliance of the cuff-arm system (C) was estimated simply as the bulk slope of the relationship over the deflation cuff pressure range of 140 to 40 mmHg. The compliance scale factor was applied to the two surrogate arm blood flow rates for two arm blood flow rate estimates in L/min (C·DO_max_·HR and C·DPVR·HR). In the third step, a second scale factor given by the ratio of the body surface area to the arm surface area (arm circumference times the cuff length) of the patient was applied to the two arm blood flow rate estimates for two CO estimates in L/min (δ·C·DO_max_·HR and δ·C·DPVR·HR).

**Fig. 5:**
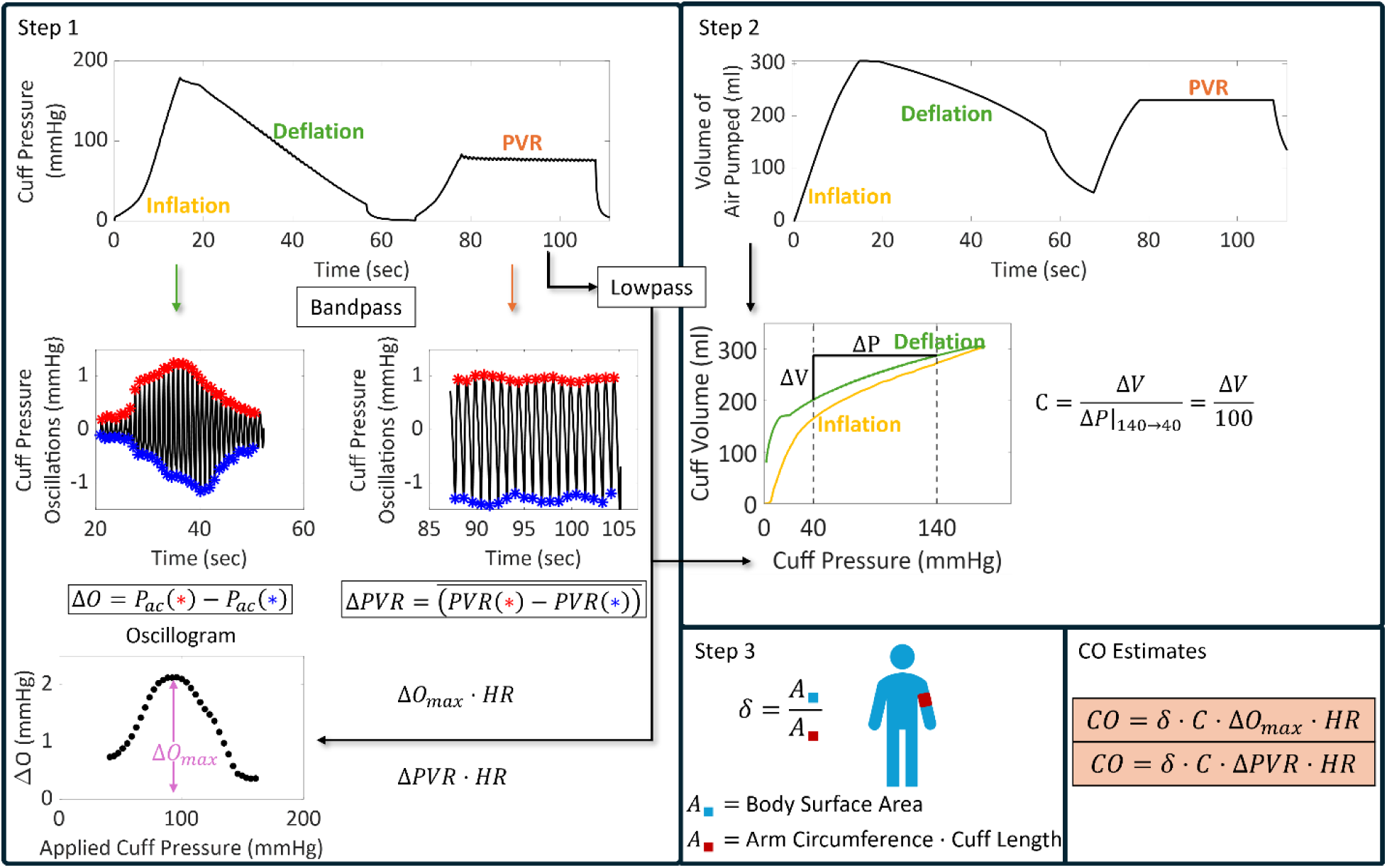
Arm cuff-based CO estimation. Two basic analyses applied to the cuff device measurements to estimate CO in L/min as the product of the (1) maximum cuff pressure oscillation amplitude during deflation (ΔO_max_) or the pulse volume recording amplitude (ΔPVR) times heart rate (HR), (2) compliance of the cuff-arm system (C), and (3) ratio of the body to arm surface area (δ).

For comparison, we applied classic pulse contour analysis to the invasive BP waveform during the deflation cuff pressure measurement. We used the Liljestrand and Zander method, which performed the best in a previous comparison of eight classic pulse contour analyses^15^. As shown in Fig. 6, this method calculated the average of SP, DP, PP, and HR over the invasive BP waveform and then estimated CO, normalized by the arterial compliance scale factor of the patient, as PP·HR/(SP+DP). Note that PP is a commonly used surrogate of stroke volume, and 1/(SP+DP) is a correction to account for the inverse relationship between arterial compliance and BP. We also combined the cuff and invasive CO estimates for a fourth CO estimate.

**Fig. 6:**
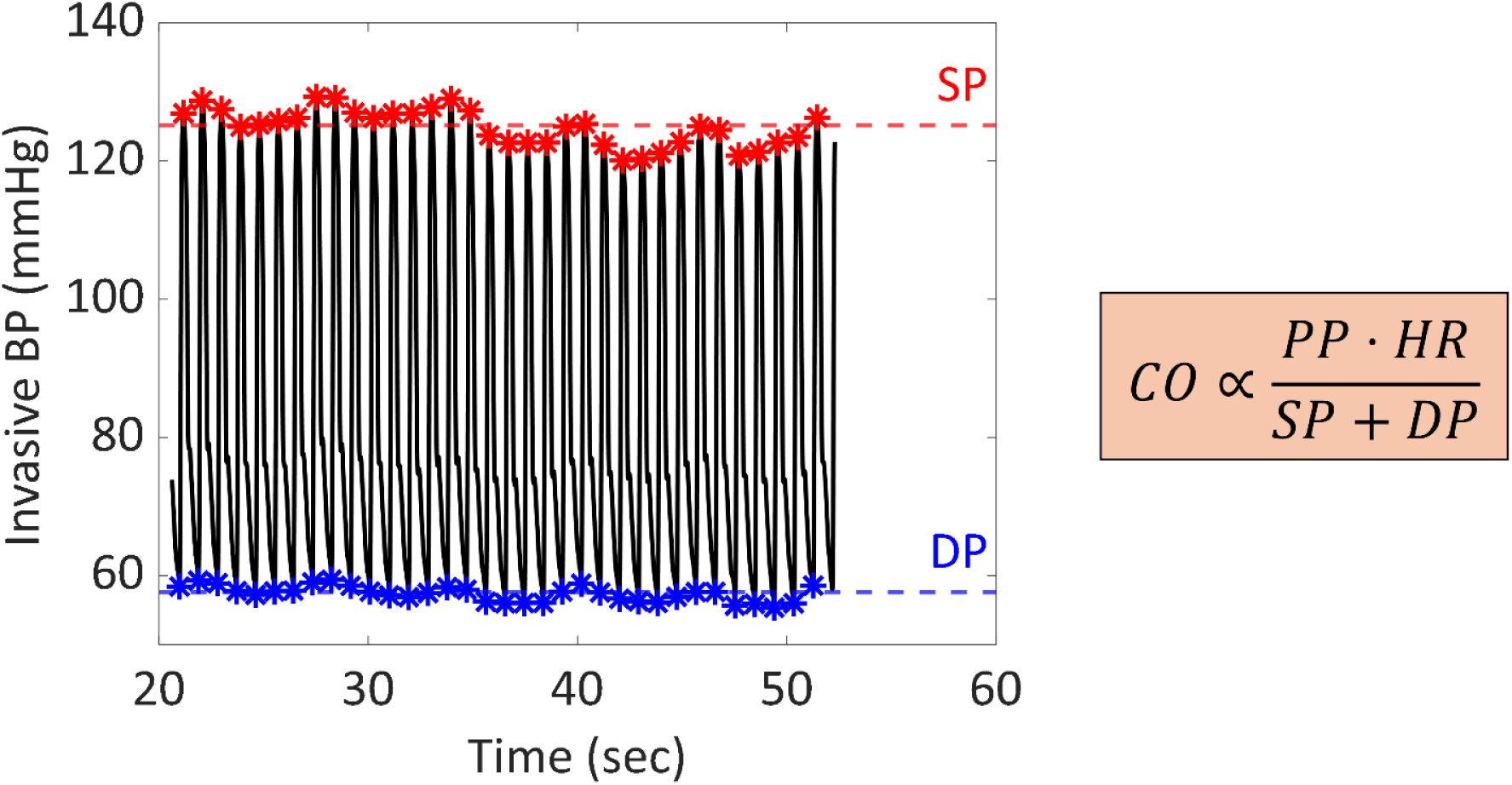
Invasive pulse contour CO estimation for comparison. Classic pulse contour analysis applied to the invasive BP waveform to estimate CO normalized by arterial compliance scale factor of the patient as pulse pressure (PP) times HR divided by the sum of systolic and diastolic BP (SP+DP)^15^.

We averaged each the four CO estimates as well as each of the two arm blood flow rate surrogates and two arm blood flow rate estimates over the two inflation-deflation cycles. Similarly, we averaged the corresponding bolus thermodilution CO values. We evaluated each of the eight estimates against the reference CO using correlation analysis. Note that we did not employ Bland-Altman analysis, because six of the estimates were not of absolute CO in L/min. We also evaluated the estimated relative CO changes between consecutive measurements (ΔCO=100·(CO_after_-CO_before_)/CO_before_)) in individual patients against the reference CO changes for directional accuracy using standard concordance analysis with a 15% exclusion zone. We lastly evaluated the estimated relative CO changes against the reference CO changes for agreement using Bland-Altman analysis and the overall root-mean-square-error metric (= √(µ^2^+σ^2^), where µ is the Bland-Altman bias error and σ is the Bland-Altman precision error). Note that the popular percentage error metric (i.e., twice the precision error divided by the grand mean of the device and reference CO) is actually a flawed metric that rewards overestimation of the CO^3^.

## Results

Table 1 summarizes the patient data included for analysis. The data comprised a total of 191 sets of cuff device measurements, the invasive BP waveform, and reference CO from 24 liver transplant and 10 cardiac surgery patients. As expected, the average CO was high for liver transplant patients but low for the cardiac surgery patients, and the CO changed appreciably, especially in the liver transplant patients. CO changes due to vasopressor therapy were captured more than due to fluid therapy in the patient cohort.

**Table 1:** Summary of patient data for analysis.

| **Variables** | **Liver Transplant** | **Cardiac Surgery** |
| --- | --- | --- |
| Number of Patients, n (%) | 24, (71%) | 10, (29%) |
| Number of Measurement Sets, m (%) | 164, (86%) | 27, (14%) |
| Age (years) | 52 ± 15 | 68 ± 13 |
| Male/Female, n-n (%-%) | 14/10 (58-42%) | 6/4 (60-40%) |
| Height (cm) | 173 ± 9 | 173 ± 13 |
| Weight (kg) | 87 ± 17 | 89 ± 26 |
| Body Surface Area (m<sup>2</sup>) | 1.9 ± 0.2 | 1.9 ± 0.3 |
| Arm Circumference (cm) | 28 ± 3 | 28 ± 3 |
| CO (L/min) | 9.3 ± 2.8 | 4.3 ± 1.3 |
| Within Patient CO SD (L/min) | 1.6 ± 0.8 | 0.7 ± 0.6 |
| HR (bpm) | 88 ± 13 | 74 ± 13 |
| PP (mmHg) | 70 ± 16 | 67 ± 21 |
| SP (mmHg) | 124 ± 19 | 126 ± 27 |
| DP (mmHg) | 54 ± 10 | 59 ± 14 |
| Fluid Therapy, m | 6 | 0 |
| Vasopressor Therapy, m | 54 | 6 |

Values are mean±SD. CO is cardiac output; SD, standard deviation; HR, heart rate; PP, pulse pressure; SP, systolic blood pressure; DP, diastolic blood pressure.

The CO estimates via analysis of the cuff device measurements during the PVR were not as accurate as those obtained during the deflation (Fig. 5). This finding was maintained even after attempting different methods to compute the compliance of the cuff-arm system (C) such as using the cuff volume-pressure relationship during inflation to the PVR. The PVR-based results are therefore not presented below.

Fig. 7 shows correlation plots of the estimates at each of the three major steps of the analysis of the cuff device measurements during deflation (Fig. 5) versus the reference CO. The surrogate of arm blood flow rate in mmHg/min (ΔO_max_·HR) from the first step yielded a correlation of 0.42 with the reference CO (Fig. 7A). The arm blood flow rate estimate in L/min (C·DO_max_·HR) from the second step produced an appreciably higher correlation of 0.57 (Fig. 7B). The final CO estimate in L/min (δ·C·DO_max_·HR) from the third step resulted in a mildly higher correlation of 0.60 but was more importantly distributed around the identity line (Fig. 7C). The root-mean-square-error was 2.78 L/min. The figure also illustrates notably improved intra-patient correlation via the C scale factor in a case in which the cuff was changed from one size (green circles) to another (red circles) in the middle of liver transplantation (Fig. 7ABC).

**Fig. 7:**
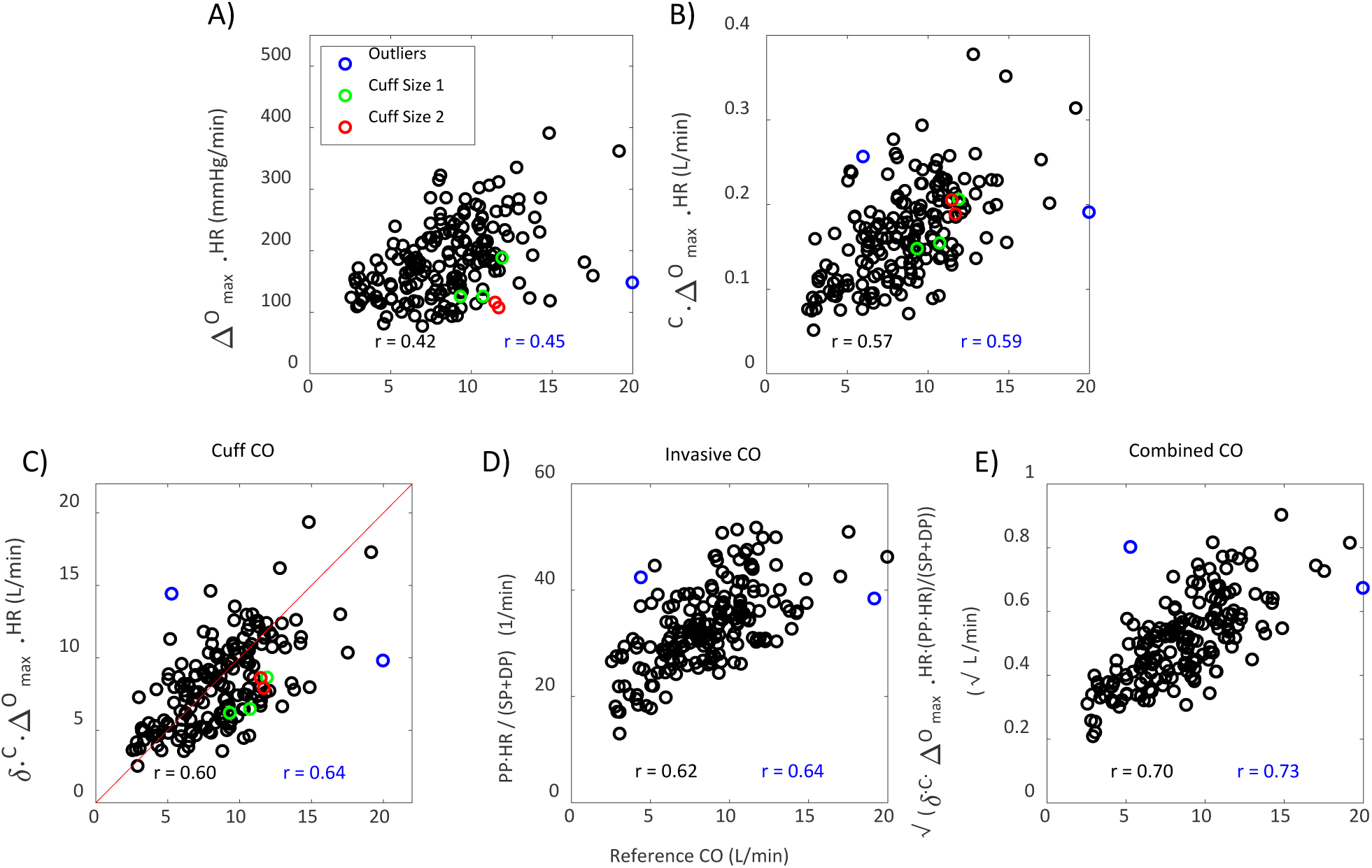
Correlation plots of the estimates via the deflation cuff device measurements at each of the three analysis steps, classic invasive pulse contour analysis, and the geometric mean of the cuff and invasive CO estimates versus the reference CO. Blue circles are outliers; green circles, estimates from cuff size 1 in a patient; red circles, estimates from cuff size 2 in the same patient; r, correlation coefficient over all datapoints (black) and minus the outliers (blue).

Fig. 7 additionally shows correlation plots of the CO estimates via classic invasive pulse contour analysis and via a combination of the cuff and invasive CO estimates versus the reference CO. The CO estimate via the invasive pulse contour analysis (PP·HR/(SP+DP)) produced a similar correlation to the cuff device of 0.62 (Fig. 7D). As expected, this CO estimate was not near the identity line. Since the CO estimate via invasive pulse contour analysis and the CO estimate via the cuff device were of different scales, we combined them using the geometric mean. This simple combination of the two CO estimates appreciably improved the correlation to 0.70 (Fig. 7E).

Fig. 8 shows concordance and Bland Altman plots of the relative changes in the three CO estimates (in Fig. 7) between consecutive measurements of individual patients versus the reference CO changes. The estimated relative CO changes via analysis of the cuff device measurements during deflation yielded a concordance rate of 83% and a root-mean-square-error of 30% against the reference CO changes (Fig. 8A). Note that the compliance of the cuff-arm system (C) did not change much in each patient, because the cuff was kept in place on the arm in all patients but one, and using this scale factor did not markedly improve the concordance rate (from 78% to 83%; not shown). For comparison, the estimated relative CO changes via the classic invasive pulse contour analysis produced a mildly lower concordance rate of 81% but a lower root-mean-square-error of 26% (Fig. 8B). The estimated relative CO changes of the invasive pulse contour analysis tended to be compressed compared to the reference CO changes, whereas the corresponding changes of the cuff device showed more scatter versus the reference CO changes about the identity line (compare Fig. 8A to 8B). The estimated relative CO changes via the geometric mean of the two estimates increased the concordance rate to 85% and lowered the root-mean-square-error to 24% (Fig. 8C).

**Fig. 8:**
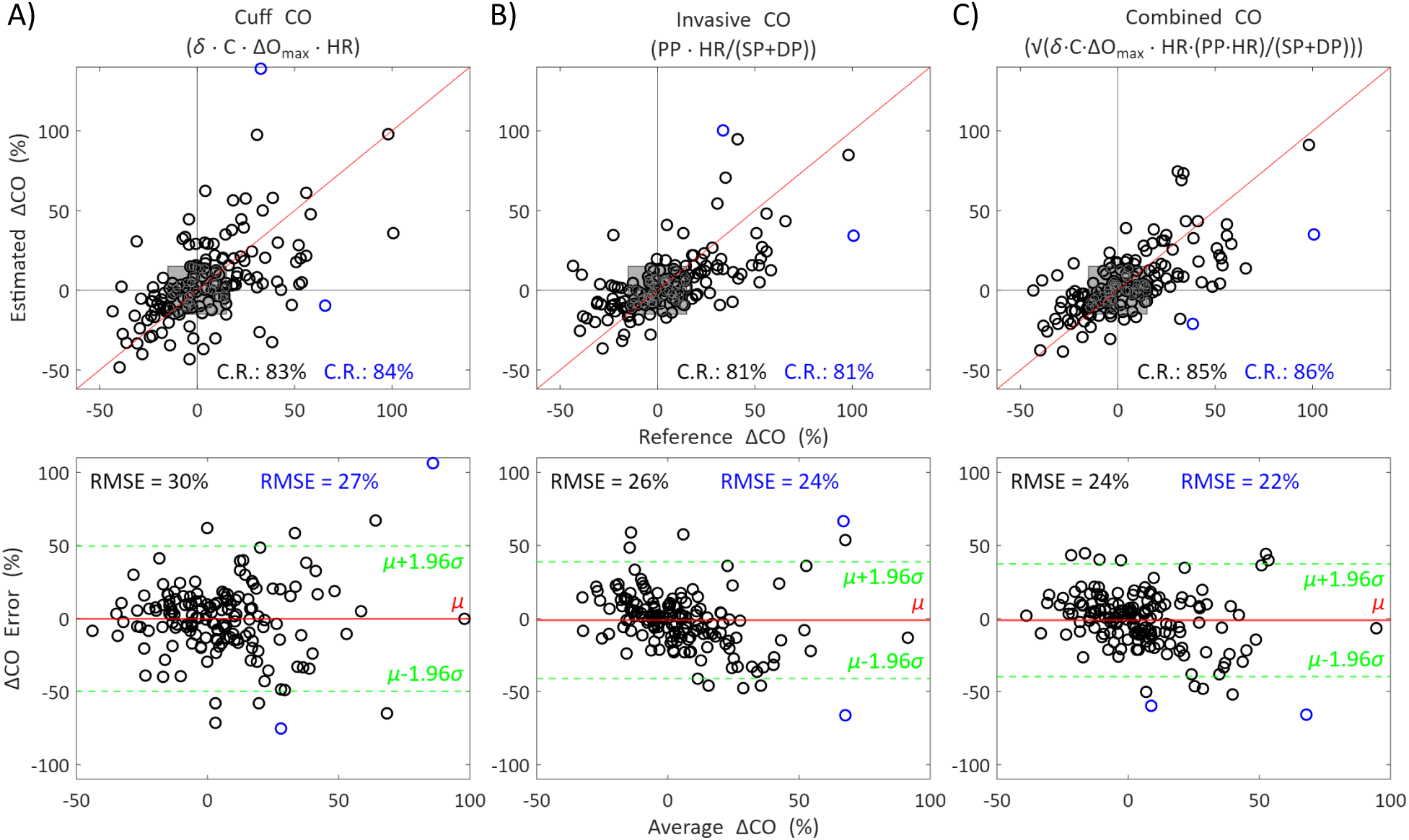
Concordance and Bland-Altman plots of the estimated relative CO changes between consecutive measurements in a patient versus the reference CO changes. Blue circles are outliers; C.R., concordance rate; RMSE, root-mean-square-error (=√(µ^2^+σ^2^), where µ and σ are the mean and SD of the errors) over all data (black) and minus the indicated outliers (blue).

There were a couple of outliers for each estimate. When these datapoints were removed, the accuracy metrics improved noticeably (blue circles in Figs. 7 and 8).

## Discussion

In this study of liver transplant and cardiac surgery patients, we found that CO monitoring may be feasible using automatic oscillometric arm cuff technology. CO monitoring can facilitate hemodynamic management of surgical and intensive care patients. CO trending in particular can help identify the cause of incipient hypotension and guide fluid, vasopressor, and inotropic therapies. Several CO monitoring devices are currently available including the pulmonary artery catheter, invasive pulse contour devices, non-invasive (volume-clamp) pulse contour devices, transthoracic and transesophageal ultrasound, and impedance cardiography^16^. However, these devices are invasive, require a skilled operator, or employ special instruments. As a result, none are widely used for patient care today. By contrast, oscillometric arm cuff devices are non-invasive, automatic, and standard instruments employed for BP monitoring in surgical and intensive care patients. Moreover, the cuff pressure oscillations that these devices measure are directly related to pulsatile blood volume rather than BP.

We used our custom cuff device, which makes a unique measurement of the volume of air pumped into and out of the cuff and measures the cuff pressure during servo-controlled linear deflation followed by a constant cuff pressure to yield a high amplitude PVR on average. We obtained measurements with this device, the invasive radial BP waveform, and reference pulmonary artery catheter-bolus thermodilution CO from 34 liver transplant and cardiac surgery patients during major surgical landmarks and following hemodynamic therapies. The reference CO varied widely across patients and changed appreciably within patients. The patient data therefore offered an interesting platform for initial testing of our hypothesis. We applied basic analysis to the cuff device measurements to estimate CO without using any training data. Our CO formula of δ·C·DO_max_·HR is the product of the maximum oscillation amplitude and heart rate (ΔO_max_·HR from the deflation cuff pressure measurement), the compliance of the cuff-arm system (C from the measured deflation cuff volume-pressure relationship), and the body to arm surface area ratio (δ from anthropometric information).

Our cuff-based method provides a measure of the arm blood flow rate, which represents only a small fraction (∼2%) of the CO^17^. However, the arm blood flow rate may change similarly to CO in response to typical interventions during surgery and intensive care. Our method also does not account for the fraction of the blood flow that directly passes through the compliant brachial artery and into the resistive lower arm arteries (“systolic runoff”)^18^. However, the method makes the measurement at zero transmural pressure (i.e., when the cuff pressure equals mean BP), a condition wherein the brachial artery compliance is maximal to minimize the errors introduced by the systolic runoff.

Nevertheless, our arm cuff-based method could estimate CO reasonably with a correlation of 0.60 and a concordance rate of 83% against the reference CO. Using the compliance of the cuff-arm system (C), which differs from patient to patient (e.g., Fig. 1), to convert the cuff pressure oscillations in mmHg to blood volume oscillations in ml improved the correlation appreciably. Furthermore, this compliance was critical for CO estimation in the one patient in whom the cuff was replaced mid-surgery. Therefore, incorporating the air volume measurement in the device to compute the cuff-arm compliance proved important. Using the body to arm surface area ratio scale factor (δ) brought the estimated CO to a level similar to the reference CO. Therefore, using this obvious anthropometric scale factor to approximately calibrate the arm blood flow rate to CO proved effective. It is also worth noting that some of the discrepancy between the arm cuff-based and reference methods may be attributed to error in the reference method, as bolus thermodilution does not provide a gold standard CO measurement^19^.

Perhaps surprisingly, the accuracy of the arm cuff-based method was essentially the same as classic invasive pulse contour analysis, which yielded a correlation of 0.62 and concordance rate of 81% against the reference CO. The CO estimates of the pulse contour analysis were normalized by the patient-specific arterial compliance scale factor and therefore not of the same scale as the reference CO. A formula to approximate the patient’s arterial compliance scale factor and thereby calibrate the pulse contour estimate to L/min is not obvious. Commercial pulse contour devices use a proprietary formula built with training data to estimate the arterial compliance scale factor from the BP levels and basic anthropometric information^3^. Such formulas will surely bring the estimated CO to the same level as the reference CO but are only approximate and therefore may not necessarily improve the correlation to a significant extent. For example, although CO estimation results are not strictly comparable across different patient cohorts, commercial pulse contour devices have shown correlations of 0.5-0.8 against pulmonary artery catheter-bolus thermodilution CO in recent studies^10–13^. Therefore, the comparison of the arm cuff-based method and classic invasive pulse contour analysis in terms of correlation may be viewed as apples-to-apples in the sense that neither method used training data but as apples-to-oranges in the sense that the arm cuff-based method provided an absolute CO estimate whereas invasive pulse contour analysis provided a proportional CO estimate. However, the comparison of the two methods in terms of concordance may be viewed strictly as apples-to-apples.

Because the cuff and invasive CO estimates are entirely independent, combining them could conceivably upgrade the accuracy. We indeed found that using a simple geometric mean of the two estimates improved the correlation to 0.70 and the concordance rate to 85%. Note that a significant fraction of surgical and intensive care patients including the ones studied herein have automatic arm cuff devices and arterial catheters in place as part of routine care^20^.

We know of one current arm cuff-based device for CO monitoring (Mobil-O-Graph, IEM GmbH, Germany)^21,22^. This device estimates SP and DP from the deflation cuff pressure measurement; measures the PVR at a subsequent sub-diastolic cuff pressure; calibrates the PVR to SP and DP; and applies pulse contour analysis to the derived brachial BP waveform. The resulting estimate may reflect the CO rather than arm blood flow rate, because, for example, PP is a surrogate of stroke volume. However, the device may not measure pumped air volume, requires additional measurement time for PVR, and must approximate the arterial compliance scale factor. Furthermore, the device may have not been tested as thoroughly against pulmonary artery catheter-bolus thermodilution CO as our device (24 measurements in 24 patients versus 191 measurements in 34 patients)^22^. Most notably, this device may have not been assessed for crucial CO trending or compared to invasive pulse contour analysis.

We also investigated the PVR at a cuff pressure near the mean BP of the patient population using our custom cuff device. The peak-to-peak amplitude of the PVR could be detected more robustly than the maximum oscillation amplitude. However, use of the PVR did not translate to more accurate CO estimation in our study. One reason could be that the PVR amplitude is affected by the prevailing mean BP, whereas the maximum oscillation amplitude is at zero transmural pressure and therefore hardly impacted by mean BP. Another reason could be appreciable systolic runoff. The maximum oscillation amplitude can also be determined from conventional cuff deflation and is therefore more convenient to measure than the PVR.

Although the patient dataset for study allowed for exciting evidence of the potential for arm cuff-based CO monitoring, this dataset was not sufficient to train and test more elaborate analyses for optimizing the CO estimation accuracy including reducing the scatter in concordance plots. For example, combining the arm blood flow rate estimates with CO estimates via a derived BP waveform from the cuff device could improve accuracy. It may also be possible to measure PP variations with the PVR, which could help in CO estimation. In addition, since the cuff volume-pressure relationship is nonlinear and dynamic, the cuff pressure oscillations could be more precisely converted to blood volume oscillations through more sophisticated analysis. The calibration to CO from available information may likewise be improved. Powerful machine learning methods built on a large training dataset may be especially helpful in bringing the CO estimation accuracy to a level congruent with clinical use (e.g., near 90% concordance rate).

In conclusion, we showed that CO monitoring may be feasible with an automatic arm cuff device that measures both the cuff pressure and volume of air pumped into and out of the cuff. Such a unique device would appear like any cuff device to end-users and therefore be entirely consistent with standard care. Future investigations of arm cuff-based CO monitoring are needed. Collection of a large set of training data comprising cuff device and reference pulmonary artery catheter-bolus thermodilution CO measurements and development of machine learning methods using this dataset are most important to maximize the CO estimation accuracy. If sufficient CO estimation accuracy can be achieved, subsequent effort to reduce the measurement time through real time analysis and step deflation should be made. It may also be worthwhile to pursue measurement of the pumped air volume via two pressure sensors and tubing of known resistance or of a pumped air volume surrogate via the cuff pressure inflation pattern (e.g., bulk slope). Potential clinical applications include more accurate CO monitoring and trending in high risk surgical and intensive care patients via combined analysis of the cuff device measurements and invasive BP waveform, non-invasive CO monitoring and trending in lower risk hospital patients instrumented with an arm cuff device, as well as non-invasive CO measurement in outpatients to help determine anti-hypertensive therapy.

## Research Support

This work was supported by the NIH Grant HL163691.

## Conflicts of Interest Statement

M. J., H. D, R. K., V. D., M. R. P., K. S., J-O. H., S. G. S., K. H-Q., A. M., and R. M. have pending patents on arm cuff-based hemodynamic monitoring technology. M. R. P., K. S., J-O. H., S. G. S., K. H-Q., A. M., and R. M. have an NIH grant on arm cuff-based hemodynamic monitoring technology. R. M. is Co-Founder and holds equity interests in Retia Medical Systems (White Plains, NY). All other authors declare no conflicts of interest.

## Data Availability

All data produced in the present study are available upon reasonable request to the authors.

## Notes

### Author Declarations

We studied patients undergoing liver transplantation or cardiac surgery (on-pump coronary artery bypass grafting) at our hospital under IRB approval (University of Pittsburgh STUDY21110091) and with written, informed consent from the patients.

